# The family as a health producer: household composition and health behaviours in a Southern Europe population

**DOI:** 10.1101/2024.09.25.24314352

**Authors:** Ricardo Alves, Judite Gonçalves, Julian Perelman

**Affiliations:** NOVA National School of Public Health, Public Health Research Centre, Comprehensive Health Research Center, NOVA University of Lisbon, Lisbon, Portugal; School of Public Health, Imperial College London, London, UK; NOVA School of Business and Economics, NOVA University Lisbon, Carcavelos, Portugal

**Keywords:** Health behaviours, household composition, Grossman model

## Abstract

**Objectives:** In the context of rapid transformations in family structures, understanding how household composition can affect adults’ health behaviours is crucial, particularly when considering the potential role of such close social relationships in shaping those behaviours. This paper documents how household structure relates to individual health behaviours.

**Study design:** We pooled cross-sectional data from the Portuguese National Health Interview Surveys of 2014 and 2019, covering 26,000+ households.

**Methods:** Linear and logistic regression models were used to assess the association between different household compositions (single dwelling adults, single parents, couples with or without children) and adherence to the Mediterranean diet, frequency of physical activity, likelihood of risky alcohol consumption, and smoking, distinguishing between men and women.

**Results:** People living alone and single parents were significantly less likely to adhere to the Mediterranean diet than individuals living in couple. Single dwellers had significantly higher likelihood of engaging in risky alcohol consumption or being smokers than individuals living in couple and/or with children. Analyses by gender revealed that women in couples with children were less likely to practice physical activity than women in couples without children; this difference was not observed among men.

**Conclusions:** Overall, family contexts strongly correlate with individual health behaviours, with people living alone or in single-parent households appearing at higher risk of having less healthy diets, risky alcohol consumption, and smoking. This study identifies key target groups for policies aiming to improve population health (behaviours) including, critically, single dwellers and single parent households.

**Key massages:** *What is already known on this topic:* Social relationships, particularly within the family, play a significant role in shaping individuals’ health behaviors. With rapid changes in family structures, understanding how household composition influences adult behaviors is crucial.

*What this study adds:* This study reveals that single dwellers generally exhibit unhealthier behaviors, except in the case of physical activity. Additionally, it highlights that having children tends to promote healthier lifestyles among couples.

*How this study might affect research, practice, or policy:* The findings underscore the importance of considering household composition in health interventions and policy development. Specifically, attention may need to be directed towards individuals living alone and single-parent households to address potential health disparities and promote healthier behaviors.

## Background

Poor diets, low physical activity, smoking, and alcohol consumption are risk factors that represent massive and growing contributions to the global burden of disease (2). Individual health is strongly determined by such behaviours, which in turn relate to close contextual factors, such as living conditions, social environment, and family structures, which have been rapidly changing. For example, in the last decade, single adult households in Europe increased in proportion of all households by 29.6%, and the average number of people per household has dropped by about 0.4 members (3). These changes may affect the relationships and dynamics within households, hence affecting behaviours and lifestyles, including those related to health. This study documents the relationship between household composition and a comprehensive array of health behaviours.

The Grossman model of demand for health has often been used to understand individuals’ health-related behaviours (4). The model posits that individual demand for medical services, or time and effort invested in healthy behaviours, result from the demand for “good health”, taking into account the trade-off between present costs and future utility. One limitation of the Grossman model, identified by Jacobson (2000), is that it is based on the individual as the sole producer of health, neglecting the influence of other family members (5). Jacobson’s proposal of the “family as health producer” model postulates that other family members’ actions and behaviours, as well as living arrangements and other household characteristics, influence the individual’s stock of health, health preferences, and behaviours.

Empirical findings mostly support that premise. Regarding diet, home-cooked and family meals tend to include more fruits and vegetables and be healthier (6,7). However, single-headed families may face obstacles to having regular family meals and spending time on food preparation at home (8–10), due to higher opportunity costs of working fewer hours with a single breadwinner, and lack of economies of scale in meal preparation costs (conceptual framework in Figure 1). Nevertheless, when there are children, these hurdles may be balanced by considerations regarding the positive externalities of healthier meals for the children. This externality argument equally applies to double-headed households, which additionally benefit from economies of scale and lower opportunity costs of meal preparation arising from task sharing at home and as breadwinners. From an empirical perspective, some studies also found that diet patterns from single or double-headed households do not significantly vary in diet quality (9,11). People living alone have no economies of scale, on the one hand, but on the other hand, may face lower opportunity costs since they do not have to devote time to childcare.

**Figure 1.**
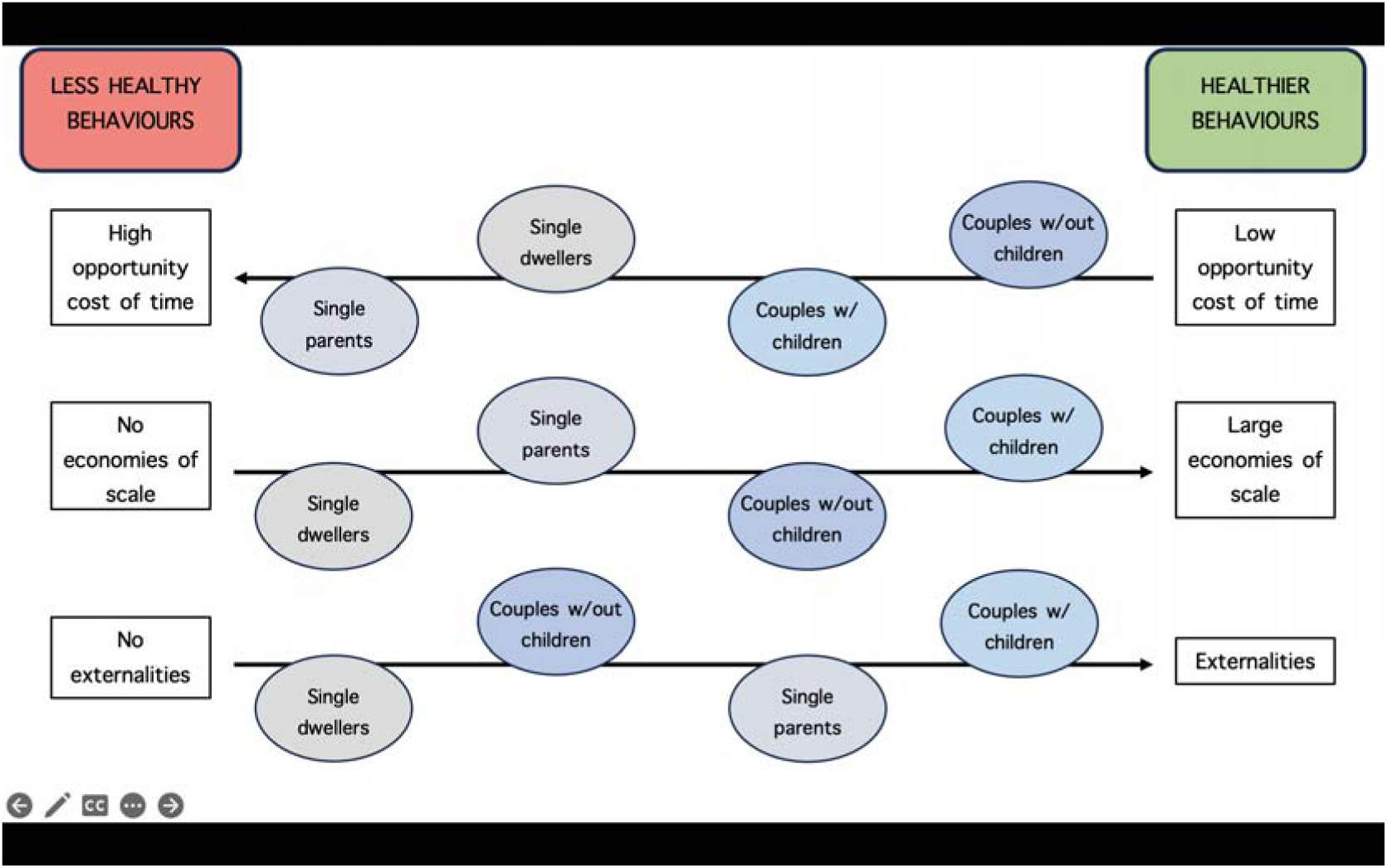
Conceptual framework: economic justifications for adoption of (un)healthy behaviours, by household structure

Family composition also plays a role in tobacco use, exercise, and risky alcohol consumption. A large meta-analysis showed the positive role of family support on exercise behaviour (12). However, adults with young children may exercise less due to lack of time caused by parental responsibilities (i.e., opportunity costs), especially when they are single parents (13). A large body of evidence indicates that adolescents are more likely to smoke when their parents also smoke, regardless of socioeconomic position (14,15). Moreover, the strongest predictors of alcohol use among adolescents are their perceptions of their parents’ drinking habits and fathers’ actual drinking (16). Childless adults may be less likely to adopt healthy behaviours due the absence of positive externalities (i.e., less worried about being a bad influence). Inversely, adults with children and, to lesser extent, childless couples, could be more prone to adopt healthy behaviours because of altruism, i.e., they account for the wellbeing benefits of them not smoking/drinking for their children and/or spouse.

Considering that each individual plays a role in the production not only of their own health, but also the health of family members, through their behaviours, our objective is to investigate the association between family structure and individual health behaviours (diet, exercise, alcohol consumption, and smoking). Given the ongoing changes in modern families’ structure, daily routines, and division of labour within the family (3,9,17), our contribution is furthered, as we provide up-to-date findings for a comprehensive range of health behaviours. Such information is relevant to inform the need and targeting of policies and interventions aiming to promote healthier behaviours and improve population health, from school or workplace programs to wider welfare policies.

Portugal is a relevant case study for both its unique features, on the one side, and its similarities to other (European) countries, on the other. Average household size and structure closely resembles the European average (3). Like many other European countries, Portugal is witnessing a rise in single-person households, and a decrease in the average number of individuals per household (3). Yet, conversely to the broader European context, but perhaps similarly to other Mediterranean countries, Portugal’s socio-cultural background entails specific gender roles in both work and family life (18,19). Despite high levels of labour market participation by both men and women, a traditional “division” of domestic and caregiving responsibilities persists, with the burden falling mostly on women. Our analyses by gender shed light on the relationship between such couple dynamics and health behaviours.

## Methods

### Study design and population

This is an observational study. We pooled cross-sectional data from the Portuguese National Health Interview Surveys of 2014 and 2019. Both surveys are based on representative samples of the non-institutionalized adult population living in Portugal (1,20). The surveys followed a regional and multistage stratified sampling scheme. The primary units (areas) were systematically selected in proportionality to the number of households in the region, and the secondary units (households) were based on random sampling within primary units. The data were collected by the National Institute for Statistics and are available on demand. This survey followed ethical standards, approved by the appropriate ethics committee, and complied with the Declaration of Helsinki. Patients and the public were not and will not be involved in the design, or conduct, or reporting, or dissemination plans of the research.

Data from the 2014 survey were collected between September and December 2014 and data from the 2019 survey between September 2019 and January 2020 (i.e., pre-COVID-19), through CAPI/CAWI, with one selected resident from each household. A total of 18,204 individuals were interviewed in 2014, and 14,617 different individuals in 2019 (i.e., repeated cross-sections). Similar questions and survey methodology allowed us to pool data from the two surveys.

Of the 32,821 individuals, we included only 26,464 between the ages of 25 and 79. At older ages, more people are institutionalized, and the sample loses representativeness. Moreover, older individuals tend to have less agency with regards to health behaviours, due to cognitive or health reasons. Additionally, in the Survey, the classification of households into households with or without children adopts a cut-off age of 25 years to define children. For this reason, we excluded individuals younger than 25.

### Outcomes

Adherence to the Mediterranean Diet (MD) was assessed using an adapted version of the KIDMED index (21,22). Our adapted index considered 13 different diet-related questions. Survey participants were asked if they had eaten the following types of foods in the previous day: legumes, fish, fruit, fresh fruit juice, vegetables, vegetable-based soup, bread, potatoes, pasta or rice, and dairy products (nine common foods in the MD), sweets, sugar-sweetened beverages, fast food, and meat (not representative of the MD). Foods characteristic to the MD were scored +1, while the four less healthy types of food were scored -1. Compared to the original KIDMED index, our modified version excluded 3 types of food: olive oil, pulses, and baked goods or pastries, because this information is not available in the Survey. As a result, the values in our adjusted index span from strong adherence (>6), moderate adherence (3-5), to weak adherence (<2), departing from the initial three-tier scale: strong adherence (>8), moderate adherence (4-7), and weak adherence (<3).

The frequency of physical activity was measured by the average number of days per week that participants engaged in exercise, including sports and leisure activities.

Risky alcohol consumption and smoking were coded as dichotomous variables. Alcohol consumption was considered risky if the participant reported risky alcohol consumption occasions (≥ 6 drinks in one episode, 10 grams of alcohol each) at least 2 - 3 days per month over the last 12 months. Available evidence suggests that frequent episodes of binge drinking may lead to accelerated alcohol metabolism and disruption of antioxidant mechanisms, leading to adverse health outcomes beyond the period of intoxication (23). Finally, participants who reported smoking daily were coded as being smokers.

### Key explanatory variable and covariates

The explanatory variable of interest was household composition (five different groups): people living alone (reference category), couples without children under 25 years old, couples with children under 25, single parents with children under 25, and other household compositions (i.e., two, non-couple, or more adults —e.g., adult with elderly parents).

Gender, age (25-39, 40-64, and 65-79), survey year (2014 and 2019), income quintiles, education (no education, primary, secondary, and tertiary education), employment status (employed, unemployed, and not in labour force —retirees, students, persons with a disability, and homemakers), and region (North, Centre, Lisbon Metropolitan Area, Alentejo, Algarve, Madeira, and the Azores) were included as covariates. The National Institute for Statistics provides only income quintiles and not income data, for confidentiality reasons.

### Statistical Analysis

Linear regression models were used to evaluate the association between different household compositions and adherence to the MD and frequency of physical activity. Risky alcohol consumption and smoking habits were analysed using logistic regressions. All regression models controlled for the covariates listed in the previous section. For ease of interpretation, results are presented in the form of adjusted means and probabilities, calculated for each type of household. Confidence intervals were computed based on standard errors adjusted for heteroskedasticity.

All analyses were also stratified by gender, taking into account previous findings from the health behaviours literature, all well as potential differences in household roles and responsibilities between men and women (9,24,25).

To check that results were not influenced by methodological differences across the 2014 and 2019 surveys, as a robustness check we also conducted stratified analysis by year.

## Results

A total of 26,464 participants were included, from both the 2014 and 2019 surveys. Table 1 presents the sample characteristics. The mean MD score was 3.8, the mean number of days of exercise per week was 1, 7% of the individuals reported risky alcohol consumption, and almost 18% were daily smokers. Around 1 in 3 participants were members of couples without children under 25 years old, 25% were members of couples with children under 25 years old, 23% were adults living alone, 5% were single parents with children under 25 years old, and other household compositions represented the remainder 15% of the total sample. The gender composition of the sample is 44% males and 56% females.

**Table 1.**
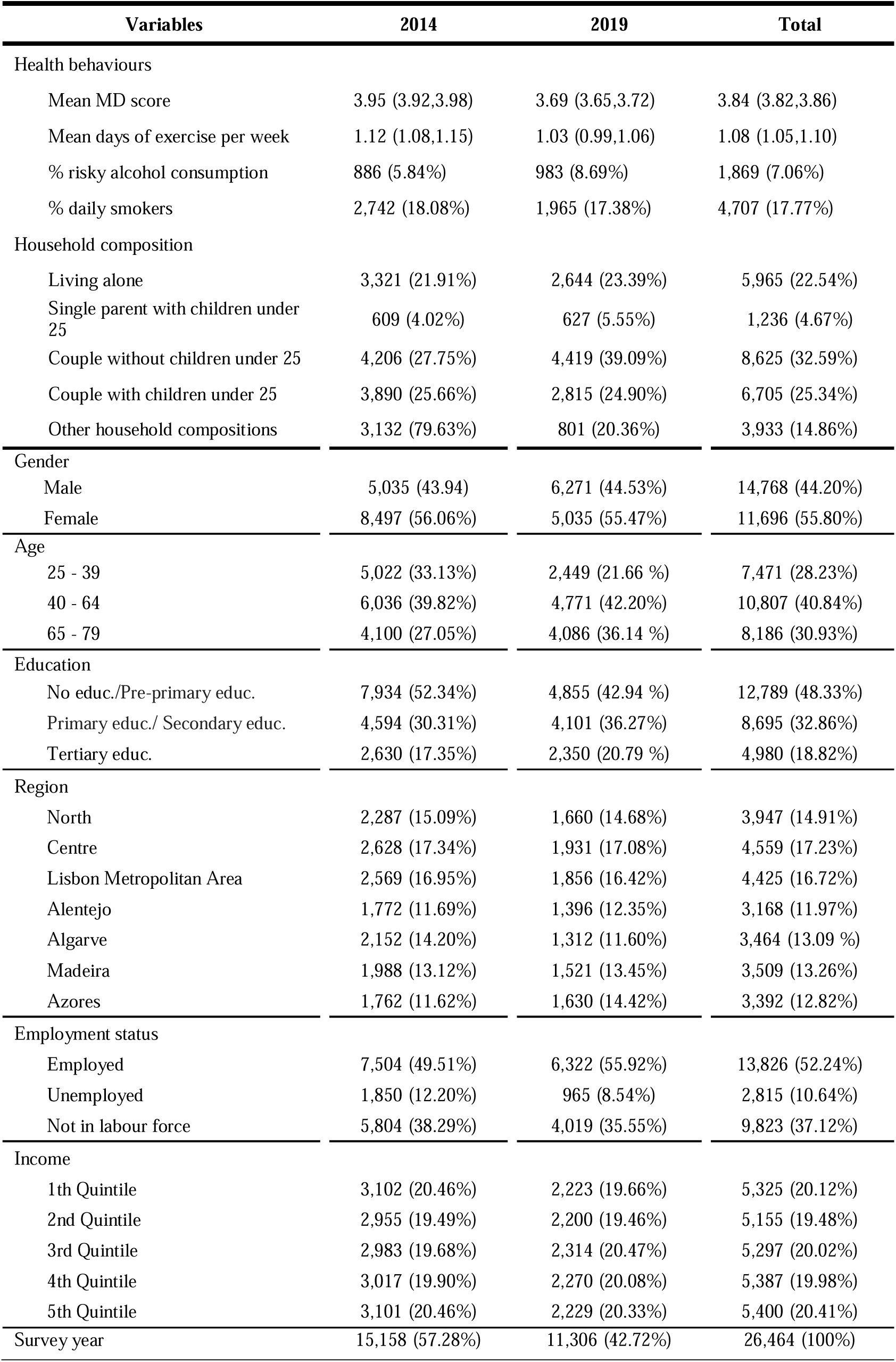
Baseline characteristics of participants in the 2014 and 2019 Portuguese National Health Surveys (95% conf. intervals for means)

The main results are presented in Table 2 and Figure 2. Focusing on statistically significant differences (P<0.05) and starting with adherence to the MD, members of couples with children (95% CI: 3.82, 3.96) and couples without children (95% CI: 3.77, 3.91) showed higher MD adherence than adults living alone, i.e., significantly higher adjusted mean MD score. The adjusted mean days of physical activity per week was significantly lower among members of couples with children (95% CI: 0.95, 1.09), compared with couples without children (95% CI: 1.15, 1.29) and adults living alone (95% CI: 1.20, 1.37). The adjusted probability of risky alcohol consumption was significantly lower among members of couples with children (95% CI: 4.3, 5.8), members of couples without children (95% CI: 4.5, 6.3), and single parents (95% CI: 3.0, 6.2), compared with adults living alone (95% CI: 7.2, 8.5). The same is true, qualitatively, for the adjusted probabilities of smoking daily: people living alone had significantly higher adjusted probability of being smokers than individuals with any other household composition (95% CI: 23.5, 27.6).

**Figure 2.**
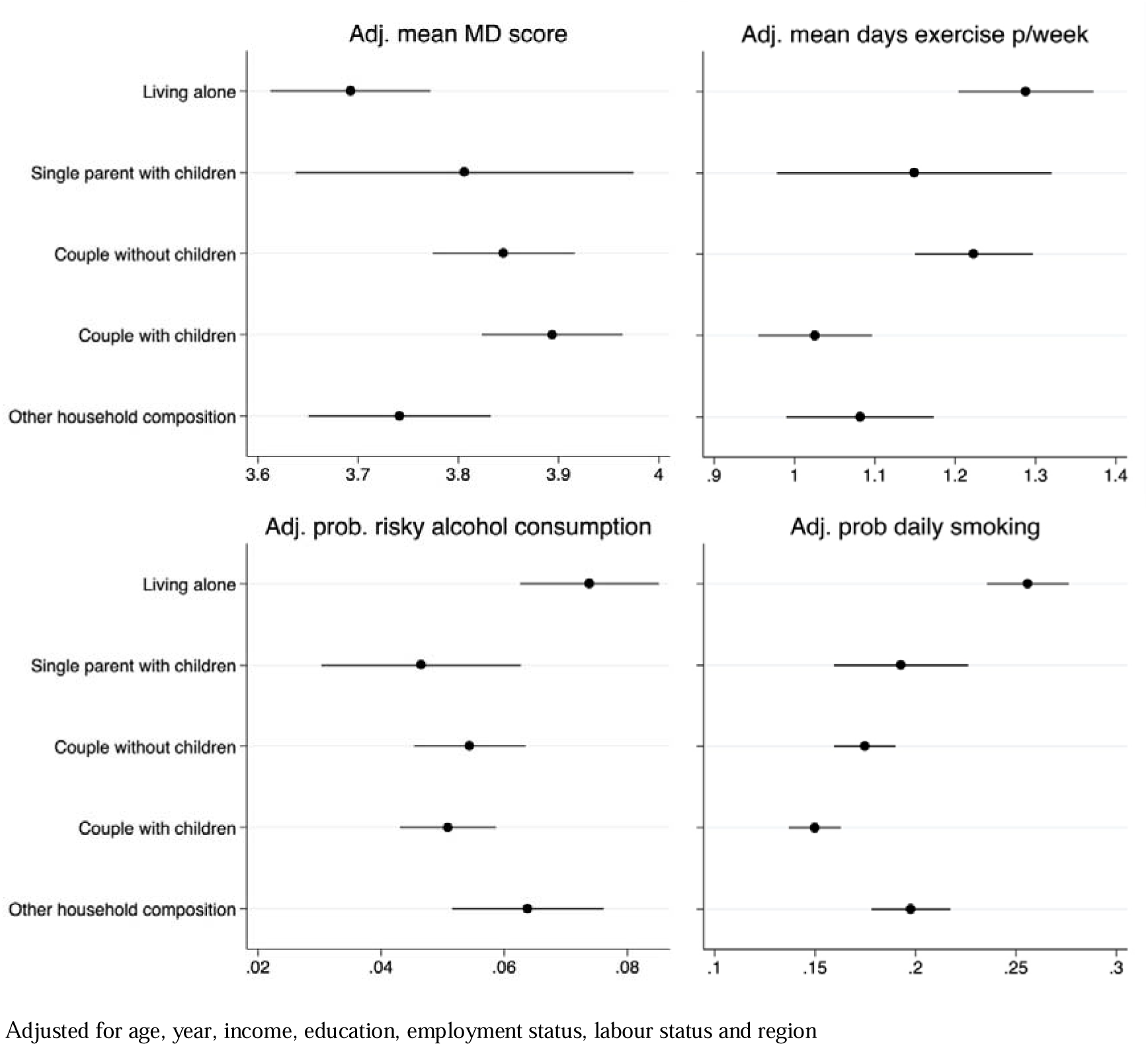
Adjusted mean MD score, mean days of exercise per week, probability of risky alcohol consumption, and probability of daily smoking by household composition (95% conf. intervals)

**Table 2.** Adjusted mean MD score, mean days of exercise per week, probability of risky alcohol consumption, and probability of daily smoking by household composition, overall and by gender (95% conf. intervals)

| Variables | Living alone<sup>Ref</sup> | Couple without children under 25 | Couple with children under 25 | Single parent with children under 25 | Other household composition |
| --- | --- | --- | --- | --- | --- |
| Adj. mean MD score | 3.69 (3.61, 3.77) | **3.84\*\***(3.77, 3.91) | **3.89\*\***(3.82, 3.96) | 3.80 (3.63, 3.97) | 3.74 (3.65, 3.83) |
| Men | 3.55 (3.42, 3.67) | 3.62 (3.52, 3.73) | 3.68 (3.58, 3.77) | 3.76 (3.32, 4.19) | 3.59 (3.46, 3.72) |
| Women | 3.83 (3.72, 3.94) | **4.05\*\***(3.96, 4.14) | **4.08\*\***(3.98, 4.18) | 3.80 (3.62, 3.98) | 3.90 (3.77, 4.02) |
| Adj. mean days exercise p/week | 1.28 (1.20, 1.37) | 1.22 (1.15, 1.29) | **1.02\*\***(0.95, 1.09) | **1.14\*** (0.97, 1.31) | **1.08\*\***(0.98, 1.17) |
| Men | 1.45 (1.30, 1.59) | 1.33 (1.22, 1.44) | **1.24\***(1.13, 1.35) | 1.36 (0.95, 1.77) | **1.21\*** (1.07, 1.35) |
| Women | 1.16 (1.06, 1.27) | 1.10 (1.00, 1.20) | **0.84\*\***(0.75, 0.93) | 1.06 (0.87, 1.24) | **0.92\*\***(0.80, 1.04) |
| Adj. prob. risky alcohol consump. | 7,3% (7.2, 8.5) | **5,4%\*\***(4.5, 6.3) | **5,0%\*\***(4.3, 5.8) | **4,6%\*\***(3.0, 6.2) | 6,3% (5.1, 7.6) |
| Men | 13,9% (11.6, 16.2) | **10,3%\*\***(8.5, 12.2) | **9,1%\*\***(7.6, 10.6) | 12,7% (6.4, 18.9) | 11,1% (9.8, 13.3) |
| Women | 1,5% (0.8, 2.1) | 1,0% (0.5, 1.4) | 1,1% (0.7, 1.5) | 1,3% (0.6, 2.1) | 1,4% (0.8, 2.1) |
| Adj. prob daily smoking | 25,5% (23.5, 27.6) | **17,4%\*\***(15.9, 18.9) | **14,4%\*\***(13.6, 16.2) | **19,2%\*\***(15.9, 22.6) | **19,7%\*\***(17.7, 21.7) |
| Men | 34,9% (31.5, 38.4) | **26,8%\*\***(24.1, 29.5) | **23,2%\*\***(20.9, 25.5) | **22,1%\*\***(13.3, 30.8) | **28,6%\*\***(25.3, 32.0) |
| Women | 17,2% (14.8, 19.5) | **9,3%\*\***(7.8, 10.9) | **7,7%\*\***(6.5, 9.0) | 16,0% (12.8, 19.3) | **11,1%\*\***(9.0, 13.2) |

Statistically significant difference in comparison with the reference category \*\* ( $p < 0.05$ ) \* ( $p < 0.10$ )
<sup>Ref</sup> Reference category
Adjusted for age, year, income, education, employment status, labour status and region

The analyses by gender reveal, first, that except for physical activity, women have healthier habits than men, with significantly higher adherence to the MD and significantly lower adjusted probabilities of risky alcohol consumption and smoking (Table 2 and Figure 3). This is true regardless of household composition, although in the case of single parents, the differences between men and women are generally not statistically significant due to lack of precision. These analyses also reveal that women exclusively drive the previous result of higher adherence to the MD among members of couples, with or without children, than among adults living alone; i.e., there are no significant differences in adherence to the MD across men living in different types of households. Similarly, less frequent physical activity among members of couples with children, compared with couples without children and adults living alone, is another result driven exclusively by women. Risky alcohol consumption among women is similarly very low across all household compositions. Among men, risky alcohol consumption is significantly less likely among members of couples with children (95% CI: 7.6, 10.6) than among single dwelling adults (95% CI: 11.6, 16.2). Lastly, single dwelling women and single mothers are significantly more likely to be smokers than women in other types of households; for men, those living alone are more likely to be smokers than those living in other kinds of arrangements.

**Figure 3.**
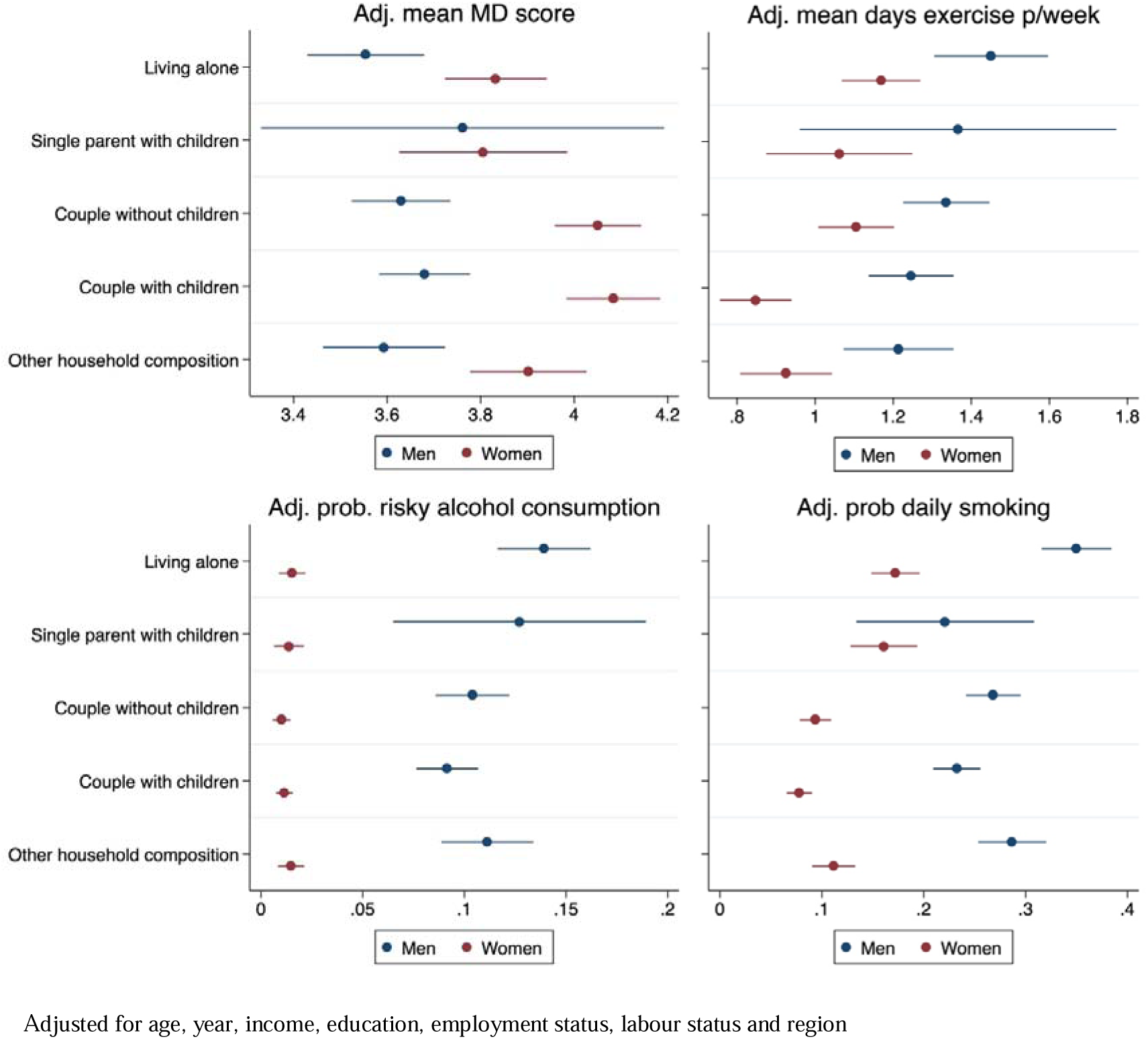
Adjusted mean MD score, mean days of exercise per week, probability of risky alcohol consumption, and probability of daily smoking by household composition, by gender (95% conf. intervals)

Our robustness check showed similar patterns in health behaviours across different household compositions in 2014 and 2019 (Table A1 in the Appendix).

## Discussion

Our study shows how family structure relates to individuals’ health-related behaviours. Generally speaking, with the exception of physical activity, the presence of a partner and the presence of children correlate with the adoption of healthier behaviours, especially among women. These results held after adjusting for economic and social characteristics, indicating that the positive externalities of family integration are, to some extent, independent from socioeconomic background.

The observed patterns align with previous scientific evidence linking close social relationships within households and adoption of healthier behaviours. Various studies highlight the role played by the interactions between family members in shaping the family’s daily routines and division of labour (26,27), with a direct impact on ability to have nutritious meals (7,28), quit smoking (29) and avoid risky drinking habits (16,30). For instance, there is evidence that dual-headed families may be in a better position to follow healthier diets because they share the cost and responsibility over the meals (7,9), and can more easily overcome everyday obstacles like lack of time to cook (8) or higher cost of some healthy foods (31). Conversely, for single-parent families, there are no economies of scale, and opportunity costs of buying and preparing food are higher because the burden is not shared. Furthermore, lack of family support has shown to result in less parental monitoring over meals and less importance or priority attached to food choices (32,33).

Prior literature suggests that spouse support may help smoking cessation (29) and that divorce increases the likelihood of smoking and binge drinking (24), which together may explain the lower likelihood of smoking and risky alcohol consumption amongst couples observed here. However, negative externalities have also been documented, i.e., smoking husband leading to wife initiating smoking (29,34). Other studies suggest a weak link between social support and smoking status (35), or hypothesize that the correlation between smoking and living with a partner reflects matching on the marriage market(36) (i.e., reverse causality). With regards to the presence of children, the lower likelihood of smoking and risky alcohol consumption may result from parents’ greater feeling of responsibility and wanting to set a good example (37). From an extended Grossman model point of view, parents may embody in their utility function not only the benefits of healthy behaviours for themselves but also for their children (i.e., altruism).

Physical activity did not follow a similar pattern to other health behaviours, suggesting a different relative importance of underlying channels (38). Lack of time (i.e., opportunity costs) due to parental responsibilities and single breadwinner status may partly explain why couples with children exercise less (13) (not smoking or not drinking does not directly require time). Sedentarism may be a less obvious “bad example” than smoking or risky drinking and may not represent a large enough incentive for a more active lifestyle. Moreover, prior literature suggests that friends, physicians, and work colleagues may play a greater role on exercise adherence than close family members (12), which could also explain why living in a dual-headed family does not seem to be particularly advantageous for physical activity.

We found that differences in MD patterns between single- and dual-headed households are only significant among women. This outcome may seem surprising, as we might expect partnered/married men to rip the most benefits from living in a dual-headed household, especially when considering the still gendered nature of home food preparation (17). However, employed men, which represent more than half of our sample, may often opt for less healthy meal choices out of home. Additionally, as women are generally more engaged in family meal planning and preparation, the positive externalities of having children may influence them more.

Lastly, the less frequent engagement in physical activity among couples with children was also only significant for women. This may reflect the unequal sharing of parental responsibilities, especially in the context of Portugal (25).

Overall, the unique socio-cultural context of Portugal with regards to gender roles, in both work and family environments, may provide an explanation for some of our findings. In Portugal, women have high labour-market participation, but still shoulder the bulk of domestic and caregiving responsibilities (18). This sets Portugal apart within the wider European context of work and family relations, as it does not conform to either a traditional male breadwinner model or a more gender equitable modern model (19). Therefore, it is important to note that the findings of this study may not be readily applicable to other European countries.

One limitation of this study is its observational design, since it draws cross-sectional data from the Portuguese National Health Surveys. So, it is not possible to follow individuals over time, to understand how changes in household structure (e.g., marriage, having children) causally impact health behaviours. Studies with a longitudinal design would be able to address endogeneity from omitted, unmeasured, variables (e.g., individual characteristics that determine both likelihood or building a family and health behaviours) or reverse causality (e.g., behaviours such as excessive alcohol consumption leading to social disconnection from family members, matching of smokers in the marriage market). Nevertheless, we don’t see this endogeneity as a major limitation, as marriage decisions and decisions to have children are hardly amenable to policy intervention. More relevant policies and interventions to promote healthy behaviours may be for example food vouchers or other welfare benefits, and our descriptive results may help identify groups that should be targeted (e.g., single parents).

One last limitation is the use of a MD-adherence index to proxy a healthy diet. This index is based on the consumption of items belonging to large food groups that do not fully account for quality or nutritional value of specific foods (e.g., fresh vs processed fish). However, more detailed dietary assessment methods like food diaries are challenging to implement on such large representative surveys. We may argue that this limitation is balanced by the representativeness of the large sample used in this study.

## Conclusion

This study found that individuals living alone and single parents may have a harder time sticking to healthy eating habits. This is likely due to factors such as high, unshared, costs and limited time for meal preparation (39,40). Positive externalities of healthy habits for other family members, especially children, may explain why smoking and risky drinking habits are less common among individuals living in couple and/or with children.

These findings are particularly relevant in the current European context, where there is a growing prevalence of single adult families (30.7% increase between 2009 and 2022) and a decreasing average number of household members (3). Although they represent a growing segment of society, single dwellers are often overlooked in political rhetoric and major policy discussions. Single dwellers are usually the sole bearers of expenses like rent or mortgage payments, utilities, and other living costs (including food), and may sometimes not have adequate social support networks.

Social security systems may play a crucial role both in the promotion of healthy behaviours, and in compensating for the detrimental impacts of unhealthy behaviours for health and social outcomes of individuals. This can be achieved through the implementation of comprehensive programs, aimed especially at single dwellers and single parent families, encompassing financial assistance (e.g., food vouchers or single family tax benefits), labour market support (e.g., inclusive employer practices, flexible work schedules, subsidized childcare) (39), and health promotion services in the community addressing adult isolation (41,42).

## Declarations

### Ethics approval and consent to participate

This survey used in this study is part of the EHIS (European Health Interview Survey) project, which regular collection is provided for in the regulations on public health statistics and health and safety at work by the European Commission (Regulation (EC) No. 1338/2008) (1). Comprehensive information about the guidelines, questionnaire, anonymization rules, ethics approval and consent procedures for participation can be found in the European Health Interview Survey section of the Eurostat website: https://ec.europa.eu/eurostat/web/microdata/european-health-interview-survey.

Consent to participate was obtained from all the participants in the study. This survey followed ethical standards, approved by the appropriate ethics committee, and complied with the Declaration of Helsinki. Patients and the public were not and will not be involved in the design, or conduct, or reporting, or dissemination plans of the research.

## Consent for publication

Not applicable

## Availability of data and materials

The data that support the findings of this study are available from Portuguese National Institute for Statistics, but restrictions apply to the availability of these data, which were used under license for the current study, and so are not publicly available. Data are however available from the authors upon reasonable request and with permission of Portuguese National Institute for Statistics.

## Authors’ contributions

RA, JG and JP contributed to the conception, design and interpretation of the work; RA also contributed to the analysis of the data. All authors read and approved the final manuscript.

## Data Availability

Availability of data and materials
The data that support the findings of this study are available from Portuguese National Institute for Statistics, but restrictions apply to the availability of these data, which were used under license for the current study, and so are not publicly available. Data are however available from the authors upon reasonable request and with permission of Portuguese National Institute for Statistics.

https://www.ine.pt/xportal/xmain?xpgid=ine_main&xpid=INE

## Acknowledgements

Not applicable

