## Supplemental material A1 for "The family as a health producer: household composition and health behaviours in a Southern Europe population": A1 Supplementary Material.docx

**Appendix**

A1. Adjusted mean MD score, mean days of exercise per week, probability of risky alcohol consumption, and probability of daily smoking by household composition, overall and by year (95% conf. intervals)

| **Variables** | **Living alone ^Ref^** | **Couple without children under 25** | **Couple with children under 25** | **Single parent with children under 25** | **Other household composition** |
| --- | --- | --- | --- | --- | --- |
| Adj. mean MD score | 3.69 (3.61, 3.77) | **3.84****(3.77, 3.91) | **3.89****(3.82, 3.96) | 3.80 (3.63, 3.97) | 3.74 (3.65, 3.83) |
| 2014 | 3.86 (3.77, 3.96) | **4.00**** (3.91, 4.08) | **4.08**** (3.99, 4.18) | 4.02 (3.83, 4.21) | 4.00 (3.91, 4.08) |
| 2019 | 3.51 (3.38, 3.64) | **3.68****(3.57, 4.68) | **3.69****(3.58, 3.79) | 3.56 (3.31, 3.80) | 3.37 (3.17, 3.56) |
| Adj. mean days exercise p/week | 1.28 (1.20, 1.37) | 1.22 (1.15, 1.29) | **1.02****(0.95, 1.09) | **1.14*** (0.97, 1.31) | **1.08****(0.98, 1.17) |
| 2014 | 1.27 (1.13, 1.39) | 1.19 (1.15, 1.31) | **1.02****(0.91, 1.14) | 1.07 (0.70, 1.21) | **1.12*** (1.07, 1.35) |
| 2019 | 1.30 (1.16, 1.43) | 1.22 (1.12, 1.33) | **1.05****(0.94, 1.16) | 1.20 (0.96, 1.44) | **0.90****(0.71, 1.10) |
| Adj. prob. risky alcohol consump. | 7,3% (7.2, 8.5) | **5,4%****(4.5, 6.3) | **5,0%****(4.3, 5.8) | **4,6%****(3.0, 6.2) | 6,3% (5.1, 7.6) |
| 2014 | 5.5% (4.3, 6.6) | 4,3% (3.3, 5.3) | **2.8%****(2.1, 3.5) | **2,1%**** (0.7, 3.5) | 4,7% (9.8, 13.3) |
| 2019 | 9,7% (7.7,11.8) | **6,9%**** (5.4, 8.4) | 8,0% (0.7, 1.5) | 7,4% (4.4, 10.4) | 7,3% (4.5, 10.6) |
| Adj. prob daily smoking | 25,5% (23.5, 27.6) | **17,4%****(15.9, 18.9) | **14,4%****(13.6, 16.2) | **19,2%****(15.9, 22.6) | **19,7%****(17.7, 21.7) |
| 2014 | 26,2% (23.7, 28.7) | **15,9%****(13.9, 17.9) | **15,2%****(13.5, 17.0) | **18,9%****(14.8, 23.0) | **19,4%****(17.3, 21.5) |
| 2019 | 25,0% (21.8, 28.3) | **19,8%****(15.9, 20.3) | **18,1%****(12.6, 16.5) | **19,8%**** (14.7, 24.9) | **19,8%****(15.6, 24.0) |

Statistically significant difference in comparison with the reference category ** (*p*< 0.05) * (*p*< 0.10)

^Ref^ Reference category

Adjusted for age, year, income, education, employment status, labour status and region
